# Indirect effects of HIV differentiated service delivery programmes on quality of clinic care: a retrospective cohort study of clients starting antiretroviral therapy

**DOI:** 10.1101/2025.09.01.25334649

**Authors:** Lara Lewis, Yukteshwar Sookrajh, Johan van der Molen, Thokozani Khubone, Lise Jamieson, Andrew Gray, Jennifer Anne Brown, Kwena Tlhaku, Francesca Little, Nigel Garrett, Jienchi Dorward, Reshma Kassanjee

## Abstract

**Introduction:** In countries with high HIV prevalence, differentiated service delivery programmes (DSD) for antiretroviral therapy (ART) play a vital role in improving access to ART for people living with HIV (PLHIV). While the benefits of DSD for clients enrolled in these programmes have been described, it is unknown whether DSD additionally benefits clients who are ineligible for these programmes, through allowing more clinic resources to be directed towards them. We aimed to assess whether increases in DSD referrals are associated with better care for clients initiating ART.

**Methods:** De-identified, routinely collected TIER.Net data from 112 clinics in KwaZulu-Natal, South Africa were used to assess service delivery for clients initiated during 2022-2023. Outcomes were the probability of newly-initiated clients having an initiation CD4 count result, a 6-month viral load result (among those visiting at 6 months) and being retained for 6 months after initiation (<90 days late for all visits). Using generalized linear mixed effects models, we measured the association of clinic DSD referral with outcomes and derived marginal probabilities for each outcome for varying DSD levels. Risk differences were calculated, with confidence intervals estimated using bootstrapping.

**Results:** Between August 2022 and October 2023, 26,226 PLHIV with a median age of 32 years were initiated on ART. Monthly DSD referral proportions at the clinics varied but increased on average from 21% in August 2022 to 29% in October 2023. Overall, 77% of clients had a CD4 count test at initiation, 72% of those attending their 6-month visit had a viral load test, and 70% were retained to 6 months. We found a positive relationship between DSD referrals and test completion, although this association was only weakly significant for viral load test; when DSD referrals increased from 20-30%, the probability of having a CD4 count and viral load test increased by 0.97% (95% CI: -0.27,2.58%) and 1.35% (95% CI: 0.11,2.95%) respectively. We found no evidence of an association with DSD referrals and 6-month retention.

**Conclusions:** In the first study of this topic, increases in DSD resulted in small improvements in care. With full DSD scale-up, however, programmatically meaningful effects could be achieved.

## Introduction

South Africa runs the largest national antiretroviral therapy (ART) programme globally, with 5.9 million people living with HIV (PLHIV) accessing the programme in 2023 alone [1, 2]. In recent years, access to ART through the national programme has been facilitated through the roll-out of differentiated service delivery (DSD) models, modes of ART delivery that are designed to make ART more accessible to PLHIV than traditional clinic-based care. The three main DSD models endorsed by the South African National Department of Health (SA NDoH), namely adherence clubs, facility-based pick-up points and out-of-facility-based pick-up points, are supported by the Central Chronic Medicines Dispensing and Distribution (CCMDD) programme [3]. In 2023, an estimated 2.6 million PLHIV in South Africa had an active CCMDD prescription [4].

The SA NDoH recommends that clients in DSD only attend clinical consultations every 6 months [5]. This is approximately half as frequent as clinical consultations for those not using DSD models which mostly occur every 2-3 months. Despite initial concerns that less frequent consultations may negatively impact the outcomes of those using DSD models, there is growing evidence that 6-monthly consultations are generally preferred by clients [6-8] and may in fact improve clinical outcomes [9-12]. Qualitative research has indicated that DSD may additionally benefit those not using DSD models, with the reduction in overall clinical consultations allowing healthcare workers to provide better quality of service to those attending clinics [13-15]. However, this has not been quantitatively measured.

Measuring the impact of the DSD roll-out on those not participating in the programme, and not only those in it, is critical to understanding the overall impact of DSD on people on ART. In this study, we explore whether the DSD roll-out has had an impact on the quality of care of PLHIV who are newly-initiated on ART in the period before which they become eligible for DSD. Specifically, we aimed to measure whether increases in the proportion of clients referred to DSD in a clinic reduced overall clinic volumes, thereby improving the outcomes of newly-initiated clients in the first 6 months on ART.

## Methods

### Study design and setting

We performed a retrospective cohort analysis using de-identified routinely collected electronic data collected between 1 February 2022 and 31 October 2023 from primary care clinics in the eThekwini municipality and uMgungundlovu districts of KwaZulu-Natal (KZN), South Africa. In 2022, the HIV prevalence in KZN was estimated to be 16% [1]. DSD programmes were introduced from mid-2016 [3], with out-of-facility-based pick-up points being the predominant mode of delivery. In this analysis, we have considered the impact of DSD programmes using out-of-facility pick-up points only because the use of other DSD models during the observation period was low. We refer to these out-of-facility pick-up point models as ‘DSD’ hereafter.

Since 2016, ART has been recommended for all PLHIV in accordance with national guidelines. Clients newly diagnosed with HIV should undergo a CD4 count test and initiate ART on the same day. Clients are required to attend clinical consultations and collect ART at their clinic approximately every 2-3 months. Once eligible, clients can be referred to DSD and collect ART from their selected pick-up point every 2-3 months, returning to the clinic for clinical consultation and a new ART prescription only every 6 months. Clients are assessed at consultations and referred back to DSD if still eligible. Therefore, DSD referral includes all clients given a new DSD script, and not just those referred to DSD for the first time. To be eligible for DSD, clients must be stable-in-care, i.e., virally suppressed, without tuberculosis, not pregnant, and without an uncontrolled chronic condition [5]. In the period used for this analysis, viral loads were measured at 6- and 12-months post ART initiation and annually thereafter [16]. Thus, newly-initiated clients could only be considered for DSD from 6 months after ART initiation.

### Data sources and data management

We analysed de-identified person-level data from TIER.Net using R 4.0 (R Foundation for Statistical Computing, Vienna, Austria) and SAS, version 9.4 (SAS Institute Inc). TIER.Net is an electronic register consisting of data on PLHIV receiving ART from South African primary care clinics [17]. It includes basic demographic data, data on visit dates to primary care clinics, referral to DSD programmes, ART start dates and regimens, prescription lengths, CD4 count and viral load measurements, pregnancy and TB screening results.

### Outcome and exposure measurement

The primary outcomes were the probability of having a CD4 count test result at ART initiation, of having a viral load test result at 6 months post ART initiation among those visiting at 6 months, and of retention for 6 months after ART initiation. For the CD4 count test outcome, we considered any CD4 count test done 6 months before or 1 month after ART initiation as completed at ART initiation. For the 6-month viral load outcome, we included clients visiting between 5 and 9 months after ART initiation, and determined whether a viral load was measured and recorded at this visit. For the retention outcome, we defined retention on ART for 6 months as not being more than 90 days late for any visit scheduled during the first 6 months on ART. Those who miss a visit were considered lost from the point of their last recorded visit.

While the outcome variables were measured on a person-level, the exposure variable was measured on a clinic-level. The exposure variable was the clinic-level proportion referred to DSD which varied over time. The proportion of clients referred to DSD will impact clinic visit attendance and healthcare worker capacity approximately 2-4 months after referral, because clients previously on 2-month dispensing referred for DSD will not require their 2- and 4-month clinic visits, and clients previously on 3-month dispensing will not require their 3-month visit. Thus, the DSD exposure variable was defined as the proportion of clients visiting in the period 2 to 4 months before the outcome measurement who are referred to DSD at any visit in the 3-month period. The value can range between 0% and 100%, although this is unlikely as a portion of visiting clients will be ineligible.

For the analysis of CD4 count and viral load tests completion proportions, we considered only one DSD exposure value, that being clinic DSD proportion measured 2-4 months prior to the outcome measurement. However, for the analysis of retention, which was measured based on clinic attendance at several time points in the first 6 months on ART, the DSD exposure variable was time-varying. The DSD exposure value used at a scheduled visit was the DSD referral proportion measured 2-4 months prior to their last attended visit, as this would have informed their decision to continue on treatment until the scheduled visit.

The analysis is restricted to using data from 2022-2023 because the DSD roll-out and clinic service delivery prior to this year were strongly impacted by the onset of the COVID-19 pandemic. We defined the DSD exposure variable using data from April 2022 onwards, to allow for the wash-out of 12-month DSD scripts that were temporarily allowed during the height of the COVID-19 pandemic [18]. Outcomes were measured from August 2022 to allow for the lag between exposure and outcome measurement. In light of this and to allow sufficient follow-up time to measure the outcomes, for the CD4 count test analysis, we included clients initiated between August 2022 and September 2023, for the viral load analysis, we included clients initiated from February 2022 to January 2023 (with 6-month visits occurring in August 2022 and July 2023 respectively) and for the retention outcome we included clients initiated between August 2022 and January 2023.

In addition to measuring a DSD exposure variable, we also defined a variable that quantifies the clinic volumes of clients seeking ART. We hypothesized that DSD impacts quality of care of newly-initiated clients through their effect on clinic volumes and, as a result, staff capacity. Developing a better understanding of the association between clinic volumes and our outcomes may therefore assist in interpreting the association measured between DSD referrals and outcomes. The variable measuring clinic volumes was defined as the number of visits to a clinic in a given month over the clinic size, where clinic size was defined as the number of clients registered to receive ART in that clinic in the given month.

### Estimand

Our estimand of interest for a given outcome was the difference in probabilities (or ‘risk’ difference) of an outcome for any two values of DSD exposure. In this analysis, we measured the risk difference when the DSD referral proportion is set to 20% compared to 10%, and when DSD is set to 30% compared to 20%, as these are commonly observed values for DSD referralS in KZN clinics based on descriptive analysis of the data.

### Confounders

Person-level variables were not considered as confounders to the association between outcomes and exposure because characteristics of newly-initiated clients, who are ineligible for DSD, are unlikely to affect DSD referrals (Supplementary Figure S1). However, certain clinic-level characteristics – such as staff capacity and skills and infrastructure– may confound the relationship: for example, better resourced clinics may have higher DSD referral proportions and provide better support to newly-initiated patients. We do not have direct measures of these characteristics, and thus these variables are considered unmeasured confounders. We did however adjust for clinics as a whole in our models, discussed below.

### Statistical analysis

We described characteristics of clients using medians, interquartile ranges (IQR), percentages and frequencies. We plotted monthly DSD referrals and outcomes across clinics over time, and the percentage of all clients on ART visiting their clinic each month against cumulative DSD referrals in the 2-4 months before.

We used generalized linear mixed effect logistic-normal models (GLMM) to regress the odds of having a CD4 count result and the odds of having a viral load result against DSD referral proportion. Model covariates included a time variable measured in months since August 2022, and clinic-specific random effects on the intercept. We included random effects to capture possible variation in outcomes by clinic, in the absence of measured clinic-level variables and to account for clustering of clients within clinics. Time was included to capture background trends in quality of care (for reasons other than the expansion of the DSD programme). In addition, covariates for year quarter of ART initiation (values 1-4), age of participant (in decades), and sex were included in models to improve precision. For the retention outcome, we used data expanded into person-month format and apply pooled GLMM to estimate the discrete-time hazard of missing a visit over a period of 1 month. Instead of including the variable measuring months since August 2022, we included a variable measuring months since ART initiation. The 6-month retention probability was derived by multiplying the complements of the estimated monthly probabilities of missing a visit over 6 months.

Next, we estimated the marginal probabilities for each outcome using numerical averaging of clinic random effects [19]. Firstly, we sampled 1000 random effects for each clinic from a normal distribution with mean vector zero and variance equal to the model-estimated variance of the random effect. Secondly, for a given DSD level, we used the realized values and the fitted fixed effect estimates to estimate 1000 conditional probabilities of the outcome and then average across the 1000 values. For the retention outcome, we obtained the 6-month probability of retention as the product of the monthly retention probabilities. Thirdly, we obtained the probability of the outcome by taking an average of the outcome probabilities above, based on the observed distribution of covariates in the study population. The three steps were repeated for varying DSD referral levels. Outcome risk differences were calculated for the contrasts 20% vs 10% and 30% vs 20%. We used nonparametric bootstrapping to obtain percentile-based 95% confidence intervals (CI) for the risk differences using 500 bootstrap samples, where sampling occurred at a clinic level.

We repeated the above model building using clinic volumes as the exposure variable.

### Ethical approval

This work was approved by University of KwaZulu-Natal Biomedical Research Ethics Committee (BE646/17), the University of Cape Town Faculty of Health Sciences Human Research Ethics Review Committee (822/2024), the KwaZulu-Natal Department of Health’s Provincial Health Research Ethics Committee (KZ_201807_021), the eThekwini Municipality Health Unit and the uMkhanyakude District Health Office and the uMgungundlovu District Health Office, with a waiver for informed consent for analysis of de-identified, routinely collected data.

## Results

Between August 2022 and October 2023, 26,226 PLHIV were initiated on ART from 112 clinics. The median (IQR) age of these clients was 32 (26-39) years, 62% were female, and most (98%) were initiated on a dolutegravir-based regimen (Table 1). External pick-points were the dominant form of delivery in these clinics and monthly referrals increased from 23% in April 2022 to 29% in October 2023. DSD referrals to external pick-up points varied across the clinics, as well as within clinics (Figure 1A). We noted that the difference in the highest and lowest monthly DSD referral of a clinic during the observation exceeded 15% in 73% of clinics. The scatterplot of monthly clinic volumes against the cumulative percentage referred to DSD in the 2-4 months prior, showed a negative trend between DSD referral and clinic volumes, with much variation in the level of clinic volumes per DSD referral level (Figure 1B).

**Table 1:** Sample description

| **Characteristics of clients initiating ART between Aug2022 –Oct2023 (N=26,226)** |  |
| --- | --- |
| Age (years), median (IQR) | 32 (26-39) |
| Female, N (%) | 16,376 (62%) |
| eThekwini district, N (%) | 17,737 (68%) |
| DTG regimen, N (%) | 25,608 (98%) |
| **Characteristics of clinics attended by newly initiated clients as of Apr2022 (N=112)** |  |
| Size (i.e., number active on ART), median (IQR) | 2,822 (1,820-4,584) |
| Proportion referred to external pick-up points, median | 21% (11%-30%) |
| Proportion referred to internal pick-up points, median | 0% (0%-3%) |
| Proportion referred to adherence clubs, median (IQR) | 0% (0%-3%) |

**Figure 1.**
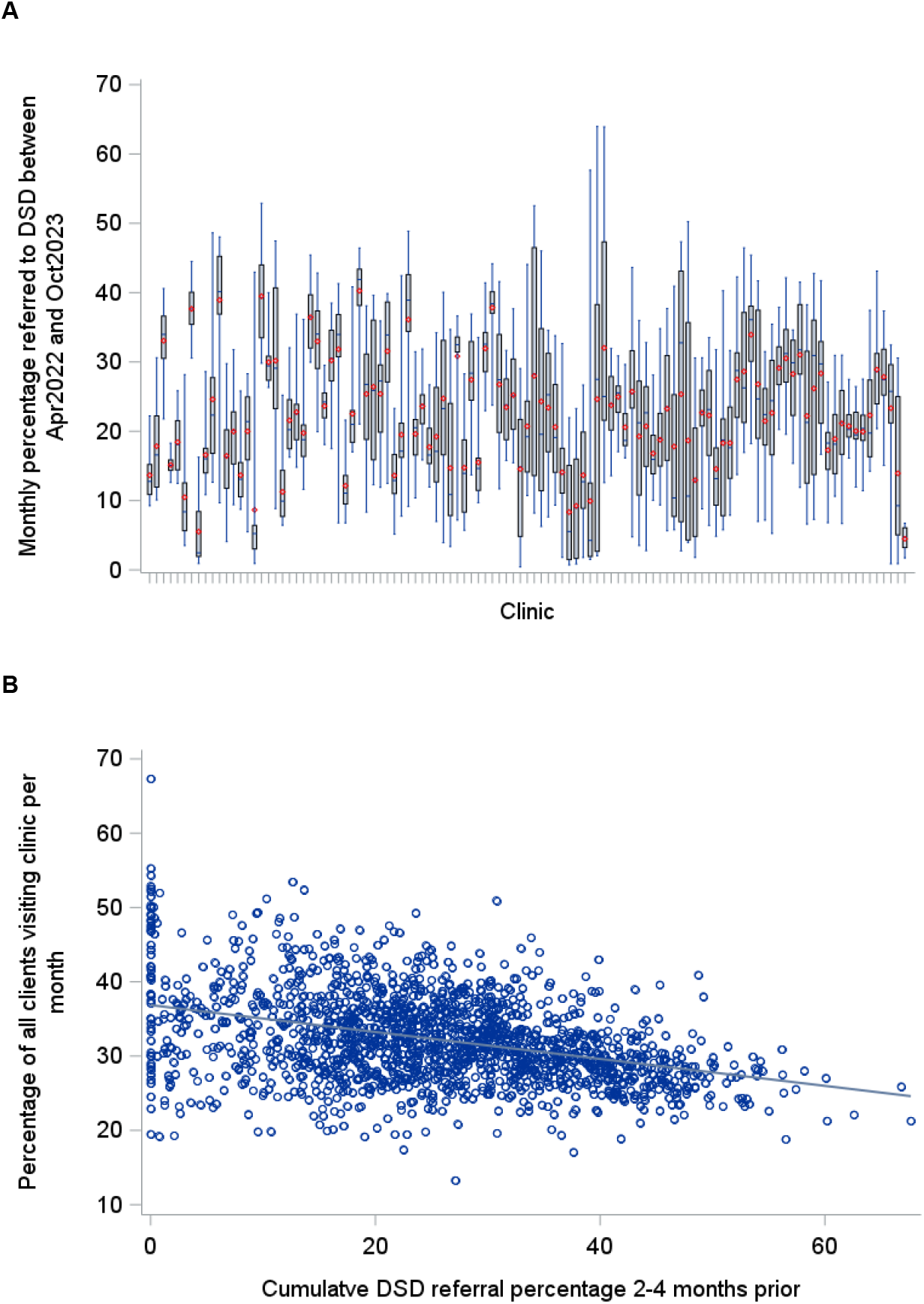
(A) Box-plot of monthly DSD referral percentages per clinic with mean values displayed in red, interquartile ranges with grey boxes, and blue whiskers extending to minimum and maximum values; (B) percentage of all clients visiting a clinic per month against DSD referral percentage 2-4 months prior, with linear regression line in grey.

The percentage of clients initiated on ART between August 2022 and September 2023 with a CD4 count test completed was 77% (19,031/24,778). Of those, most (94%) had the test performed on the same day as initiation. The CD4 count completion percentages varied by clinic (Figure 2B); the median (IQR) clinic CD4 count test completion percentage was 81% (68%-90%). Results from the GLMM suggested a positive relationship between DSD referrals and CD4 count test completion probability, although this association was small, and only weakly significant; when recent clinic DSD referrals increased from 10%-20% and from 20-30%, the probability of having a CD4 count test result increased by 0.99% (95% CI: - 0.27;2.72%) and 0.97% (95% CI: -0.27;2.58%) respectively (Table 2, Table S1). In line with these findings, we found a significantly negative relationship between clinic volumes and CD4 count test completion, with test completion probability decreasing by approximately 2% for a 10% increase in the percentage of the clinic attending a monthly clinic visit (Table 2, Table S2).

**Table 2:** Estimated marginal effect of increases in clinic DSD referrals and clinic volumes on CD4 count test completion, 6-month viral load test completion and 6-month retention among clients who were newly initiated on ART

| **Sample description** | **Outcome variable** | **#clients (#clinics)** | **Exposure variable** | **Risk difference (95% CI) change from 10% to 20%** | **Risk difference (95% CI) change from 20% to 30%** |
| --- | --- | --- | --- | --- | --- |
| Clients initiated from Aug2022 - Sept2023 | CD4 count measured | 24,778 (112) | DSD referral | 0.99% (-0.27%; 2.72%) | 0.97% (-0.27%; 2.58%) |
|  |  |  | Clinic volumes | -2.14% (-3.71%; -0.35%) | -2.27% (-4.21%; -0.35%) |
| Clients initiated from Feb2022 - Jan2023, | Viral load measured | 16,795 (112) | DSD referral | 1.38% (0.11%; 3.11%) | 1.35% (0.11%; 2.95%) |
|  |  |  | Clinic volumes | -0.94% (-2.28%; 0.67%) | -0.97% (-2.43%; 0.66%) |
| Clients initiated from Aug2022 - Jan2023 | Retained for 6 months | 11,542 (112) | DSD referral | 0.17% (-0.45%; 1.53%) | 0.17% (-0.45%; 1.48%) |
|  |  |  | Clinic volumes | 1.60% (-0.19%; 4.27%) | 1.55% (-0.19%; 3.95%) |

**Figure 2.**
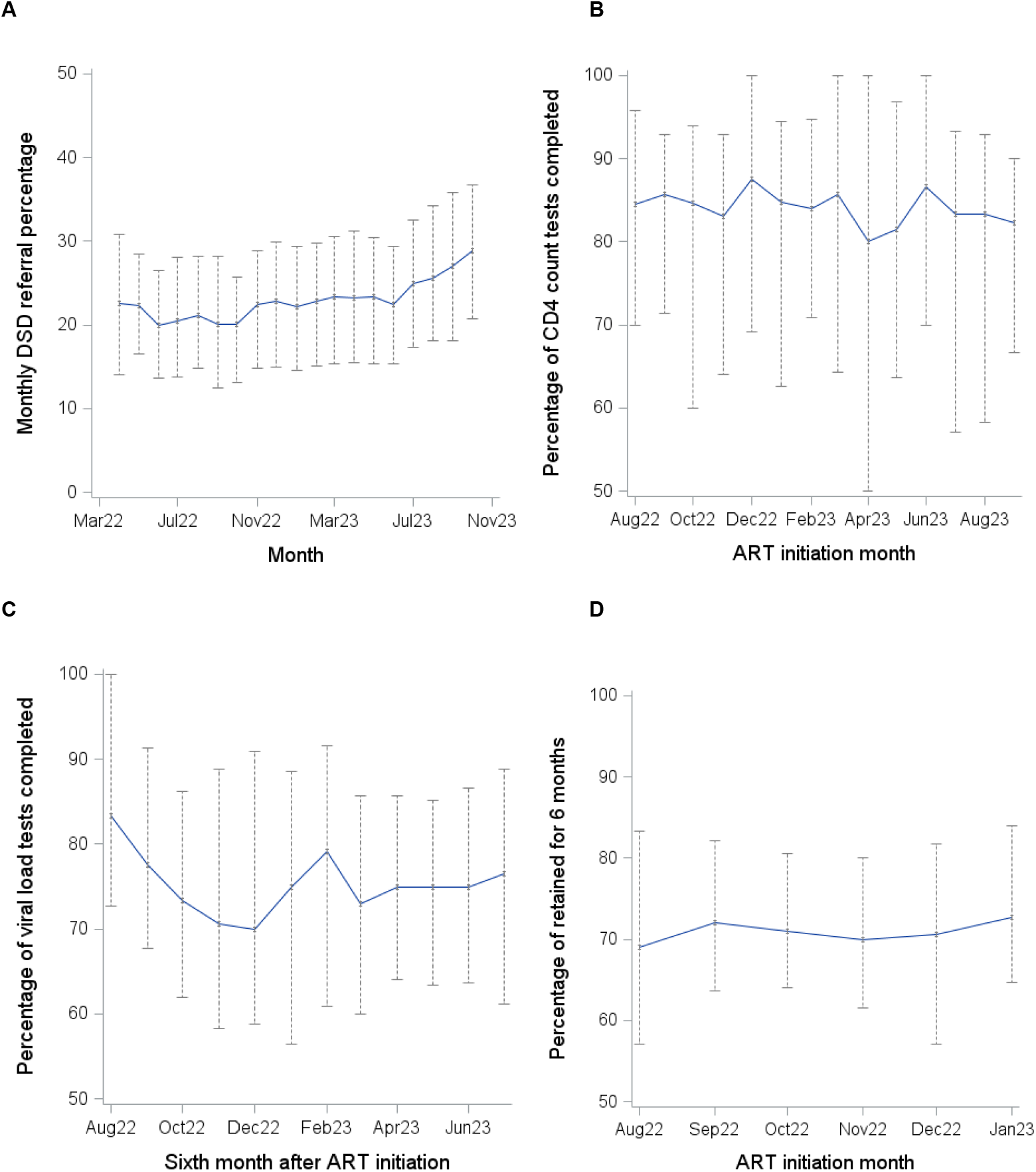
Median (blue line) and interquartile range (grey bars) of the monthly percentage of (A) visiting clients referred to DSD, (B) newly-initiated clients with CD4 count tests completed, (C) newly-initiated clients with 6-month viral load tests completed (among those attending their 6-month visit), and (D) newly-initiated clients retained for 6 months, across clinics

Among clients initiated on ART between February 2022 to January 2023 and who attended a visit between 5 and 9 months after their ART initiation, 72% (12,059/16,795) had a viral load test result. The median (IQR) time to viral load test completion from ART initiation was 188 (171-201) days. The proportion of clients with a viral load test completed varied by clinic (Figure 2C); the median (IQR) clinic viral load test completion percentage over the period was 73% (66%-83%). We found evidence of a small positive relationship between DSD referral and the probability of viral load test completion; when DSD referral prior to the visit increased from 10%-20% and from 20-30%, the probability of having a viral load test result increased by 1.38% (95% CI: 0.11;3.11%) and 1.35% (95% CI: 0.11;2.95%) respectively (Table 2, Table S1). The association between clinic volumes and the probability of having a viral load test result was negative, but not statistically significant.

Among the 11,542 clients initiated on ART between August 2022 and February 2023, 3,450 (30%) were more than 90 days late for at least one of their scheduled visits in the first 6 months. Approximately 41% of these events occurred within 1 month of ART initiation. The proportion retained for the first 6 months varied by clinic (Figure 2D). Results from the GLMM showed no evidence of an association between DSD referrals and/or clinic volumes and the 6-month probability of missing a visit (Table 2, Table S1-S2). The estimated change in 6-month retention for an increase in DSD referrals from 10%-20% and 20%-30% was 0.17% (-0.45%; 1.53%) and 0.17% (-0.45%; 1.48%) respectively.

## Discussion

In this study, we aimed to determine whether DSD has an indirect effect on the quality of care of newly-initiated clients. We analysed longitudinal data from 112 clinics, examining whether variability in each clinic’s DSD referrals over time was associated with changes in CD4 count and viral load test completion and 6-month retention of newly-initiated clients. We hypothesized that increases in referrals to DSD programmes would lower the number of clients visiting clinics, thereby allowing more resources to be allocated to new patients, improving their outcomes. Our results showed a negative relationship between clinic volumes and DSD. However, it was not perfectly linear, highlighting that there are several determinants of clinic volumes over and above DSD referrals. We found that 10% increases in clinic DSD referrals were associated with small improvements in test completion probabilities, but there was no evidence of an association with retention. Overall, these findings suggest that while early client retention may not be impacted by DSD referrals, programmatically meaningful improvements in the test completion probabilities for newly-initiated clients can be achieved with greater DSD scale-up and decanting of clients to community ART collection.

There has been little research conducted on the potential impact of DSD for ART on clients who are ineligible for DSD, against which our findings can be benchmarked. A recent study in Mozambique found that implementation of 8 different types of DSD models in the country improved population-level 12-month retention after ART initiation by 24.5% [20]. However, the use of a 12-month outcome period in this analysis, meant that the sample included a mix of clients who were enrolled in DSD, not enrolled but eligible, or ineligible, contrasting with our sample that included only those who were ineligible. In addition, although not reported, the magnitude of the DSD programme expansion in Mozambique was likely large given that eight models were utilised. The current analysis is restricted to data from 2022-2023 during which increases in clinic DSD referrals (though greater than 15% in most clinics) was unlikely to have been as large, and it is possible that the effects of reduced clinic visit frequency on retention will only be observed with greater scale-up. Recent changes to the South African ART programme, namely the widening of DSD eligibility requirements to include clients from 4 months after ART initiation [21] and the expansion of 6-month multi-month dispensing [22], are two avenues through which this can be achieved.

We chose retention as an outcome, hypothesizing that higher DSD referrals would improve client clinic experience and/or allow those in need of support more time with healthcare workers. However, particularly for clients newly initiating ART, client-specific factors, such as difficulty accepting the HIV diagnosis, ART pill burden and side effects, may outweigh or negate the benefits of improvements in clinic experiences [23]. Alternative outcome variables, including client waiting times, consultation times and the number of clients initiated on ART should also be considered for future studies. If increased staff capacity is redirected towards increasing the number of PLHIV initiated on ART rather than increasing consultation time or the time spent on processing each new initiation, the effects on quality of care per client will likely be negligible. Qualitative research to understand how additional time is prioritised within primary care clinics would be needed to complement this quantitative work.

Key strengths of our analysis include the large number of clients and clinics included in the analysis, as well as the longitudinal nature of our data. We included 26,226 newly-initiated individuals, which is just less than 20% of all new initiations in South Africa in 2023 [2]. Nevertheless, our findings must be interpreted considering the study limitations. First, confounding and/or modification by clinic characteristics, such as staff skills and quantity, infrastructure and targets, is a key concern in the analysis. We used a random effect on the intercept in regression analyses, thereby allowing for expected values of the outcomes at a given DSD level to vary by clinic. While this measure will fail to measure short-term changes in clinic qualities, the analysis was conducted over a relatively short observation period, during which large changes were unlikely. Second, the impact of possible inaccuracies in TIER.Net data also needs to be considered when interpreting these findings. Although in recent years the quality of TIER.Net data has improved, laboratory measurements captured in TIER.Net are known to be incomplete [24]. Therefore, we could not determine whether a missing laboratory test result in TIER.Net was due to missed test completion or incomplete data capture, although both are measures of service quality.

## Conclusions

This study was the first known analysis of the impact of DSD on clients initiating ART who are ineligible for DSD. Our findings suggest that modest changes in DSD referrals have a small effect on the quality of care received by these clients, but that full DSD scale-up could lead to meaningful improvements in care. Measuring the effect of DSD on other groups of ineligible clients, while it presents methodological challenges, will be important to understand the overall impact of DSD. A wider range of outcomes and larger increases in DSD referrals than observed in this study should be considered before the impact of DSD programmes on ineligible clients can be fully understood.

## Conflict of interest statement

We have no conflicts of interest to declare.

## Authors’ contributions

JD, NG, and LL conceptualised the study. YS oversaw data collection. JvdM and TK oversaw data curation. LL, JvdM, JAB and JD analysed the data, with inputs on the design and implementation from RK and FL. LL drafted the manuscript. All authors contributed to interpretation of results, critically reviewed and edited the manuscript, and consented to final publication.

## Acknowledgements

We thank the eThekwini Municipality and uMgungundlovu District Municipality Health Units as well as the staff and patients at the participating healthcare facilities.

## Funding

This work was supported, in whole or in part, by the Gates Foundation (INV-073793). The conclusions and opinions expressed in this work are those of the authors alone and shall not be attributed to the Foundation. Under the grant conditions of the Gates Foundation, a Creative Commons Attribution 4.0 Generic License has already been assigned to the Author Accepted Manuscript version that might arise from this submission. JAB is funded by the Swiss National Science Foundation (221966, to JAB). JD, Academic Clinical Lecturer (CL-2022-13-005), is funded by the UK National Institute of Health and Social Care Research (NIHR). The views expressed in this publication are those of the authors and not necessarily those of the NIHR, the National Health Service, or the UK Department of Health and Social Care.

## Data availability

The data used for this analysis cannot be shared publicly because of legal and ethical requirements regarding use of routinely collected clinical data in South Africa. The analysis code is available on request from the first author.

## Supporting Information

Supporting Information file 1: Appendix

Includes Supplementary Table 1 and Table 2 showing detailed output from GLMM used in analysis, and Supplementary Figure S1 illustrating DAGs for each of the 3 outcomes.

## Notes

### Competing Interest Statement

The authors have declared no competing interest.

### Author Declarations

Biomedical Research Ethics Committee of University of KwaZulu-Natal, the University of Cape Town Faculty of Health Sciences Human Research Ethics Review Committee of the University of Cape Town and the Provincial Health Research Ethics Committee of the KwaZulu-Natal Department of Health approved this work, with a waiver for informed consent for analysis of de-identified, routinely collected data.

