## Supplementary files for "Indirect effects of HIV differentiated service delivery programmes on quality of clinic care: a retrospective cohort study of clients starting antiretroviral therapy"

### Table S1: Full model results from generalized mixed effects models measuring association between clinic-level exposures of DSD referral proportions outcomes of CD4 count test completion, 6-month viral load test completion, and missed visit

|  |  | **CD4 count test completion** | | | **Viral load test completion** | | | **Missed visit** | | |
| --- | --- | --- | --- | --- | --- | --- | --- | --- | --- | --- |
|  |  | **Estimate** | **Standard Error** | **p-value^a^** | **Estimate** | **Standard Error** | **p-value ^a^** | **Estimate** | **Standard Error** | **p-value ^a^** |
| Intercept |  | 1.1991 | 0.1589 | <.0001 | 0.313 | 0.1256 | 0.0141 | -3.5829 | 0.1403 | <.0001 |
| DSD referral (%) |  | 0.006384 | 0.003718 | 0.086 | 0.007306 | 0.003448 | 0.0341 | -0.00075 | 0.002738 | 0.7845 |
| Age (in decades) |  | 0.02926 | 0.01664 | 0.0787 | 0.1318 | 0.01749 | <.0001 | -0.2264 | 0.01863 | <.0001 |
| Sex | female | 0.06843 | 0.03609 | 0.058 | -0.00047 | 0.0381 | 0.9902 | -0.172 | 0.03833 | <.0001 |
|  | male | 0 |  |  | 0 | . | . | 0 |  |  |
| Year quarter^b^ | 1 | 0.02405 | 0.0533 | 0.6519 | 0.0125 | 0.05282 | 0.813 | -0.08857 | 0.05259 | 0.0922 |
|  | 2 | 0.01796 | 0.06017 | 0.7654 | 0.05119 | 0.06559 | 0.4351 | n/a | n/a | n/a |
|  | 3 | 0.1009 | 0.05029 | 0.0449 | 0.248 | 0.05169 | <.0001 | 0.01262 | 0.03955 | 0.7496 |
|  | 4 | 0 |  |  | 0 | . | . | 0 |  |  |
| Time since August 2022 |  | -0.02453 | 0.004829 | <.0001 | 0.01973 | 0.006962 | 0.0046 | n/a | n/a | n/a |
| Months since ART initiation | 0 | n/a | n/a | n/a | n/a | n/a | n/a | 2.5141 | 0.1009 | <.0001 |
|  | 1 | n/a | n/a | n/a | n/a | n/a | n/a | 1.5652 | 0.1061 | <.0001 |
|  | 2 | n/a | n/a | n/a | n/a | n/a | n/a | 1.2465 | 0.1096 | <.0001 |
|  | 3 | n/a | n/a | n/a | n/a | n/a | n/a | 1.3395 | 0.1092 | <.0001 |
|  | 4 | n/a | n/a | n/a | n/a | n/a | n/a | 0.8095 | 0.117 | <.0001 |
|  | 5 | n/a | n/a | n/a | n/a | n/a | n/a | 0 | . | . |
| Variance of intercepts |  | 1.3605 | 0.1993 | <0.001 | 0.3692 | 0.06239 | <0.001 | 0.08176 | 0. 0176 | <0.001 |

^a^ *using a t-statistic for fixed effects and a mixture of chi-square distributions for the variance components; ^b^ of ART initiation for missed visit model*

### Table S2: Full model results from generalized mixed effects models measuring association between clinic volumes and outcomes of CD4 count test completion, 6-month viral load test completion, and missed visit

|  |  | **CD4 count test completion** | | | **Viral load test completion** | | | **Missed visit** | | |
| --- | --- | --- | --- | --- | --- | --- | --- | --- | --- | --- |
|  |  | **Estimate** | **Standard Error** | **p-value ^a^** | **Estimate** | **Standard Error** | **p-value ^a^** | **Estimate** | **Standard Error** | **p-value ^a^** |
| Intercept |  | 1.8755 | 0.2283 | <.0001 | 0.6682 | 0.2013 | 0.0012 | -3.3858 | 0.1795 | <.0001 |
| Clinic volumes (%) |  | -0.01533 | 0.00544 | 0.0048 | -0.0053 | 0.005264 | 0.3138 | -0.00681 | 0.004384 | 0.1205 |
| Age (in decades) |  | 0.02956 | 0.01664 | 0.0757 | 0.1317 | 0.01749 | <.0001 | -0.2285 | 0.01863 | <.0001 |
| Sex | female | 0.06813 | 0.03609 | 0.0591 | -0.00083 | 0.03809 | 0.9826 | -0.1689 | 0.03834 | <.0001 |
|  | male | 0 |  |  | 0 | . | . | 0 |  |  |
| Year quarter **^b^** | 1 | 0.02457 | 0.05331 | 0.6449 | 0.01233 | 0.05293 | 0.8157 | -0.09086 | 0.0526 | 0.0841 |
|  | 2 | 0.03491 | 0.06038 | 0.5631 | 0.0593 | 0.0661 | 0.3697 | n/a | n/a | n/a |
|  | 3 | 0.1441 | 0.05131 | 0.005 | 0.2703 | 0.05332 | <.0001 | 0.02848 | 0.04115 | 0.4888 |
|  | 4 | 0 |  |  | 0 | . | . | 0 |  |  |
| Time since August 2022 |  | -0.02959 | 0.005252 | <.0001 | 0.01954 | 0.007281 | 0.0073 | n/a | n/a | n/a |
| Months since ART initiation | 0 | n/a | n/a | n/a | n/a | n/a | n/a | 2.531 | 0.1013 | <.0001 |
|  | 1 | n/a | n/a | n/a | n/a | n/a | n/a | 1.5721 | 0.1062 | <.0001 |
|  | 2 | n/a | n/a | n/a | n/a | n/a | n/a | 1.2485 | 0.1096 | <.0001 |
|  | 3 | n/a | n/a | n/a | n/a | n/a | n/a | 1.3355 | 0.1092 | <.0001 |
|  | 4 | n/a | n/a | n/a | n/a | n/a | n/a | 0.8043 | 0.117 | <.0001 |
|  | 5 | n/a | n/a | n/a | n/a | n/a | n/a | 0 |  |  |
| Variance of intercepts |  | 1.3557 | 0.1979 | <0.001 | 0.3586 | 0.06023 | <0.001 | 0.08142 | 0.01758 | <0.001 |

**^a^** *using a t-statistic for fixed effects and a mixture of chi-square distributions for the variance components;* **^b^** *of ART initiation for missed visit model*

Figure S1: Directed acyclic graph (DAG) illustrating (A) the relationship between clinic characteristics, DSD referrals in preceding months and CD4 count test completion among clients initiating ART; (B) the relationship between clinic characteristics, DSD referrals and viral load test completion for clients who are attending a visit 6 months after ART initiation; (C) the relationship between clinic characteristics, DSD referrals and 1-month retention for clients attending a visit at month k (k=0,1,..5) after ART initiation. Clinic volumes shown as mediator.

**A**

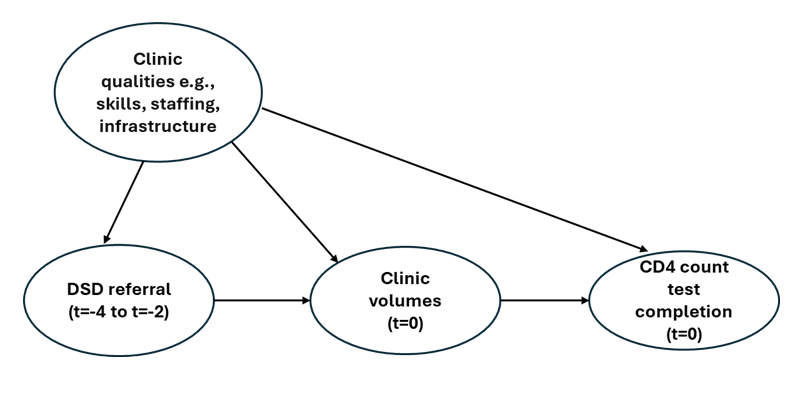

**B**

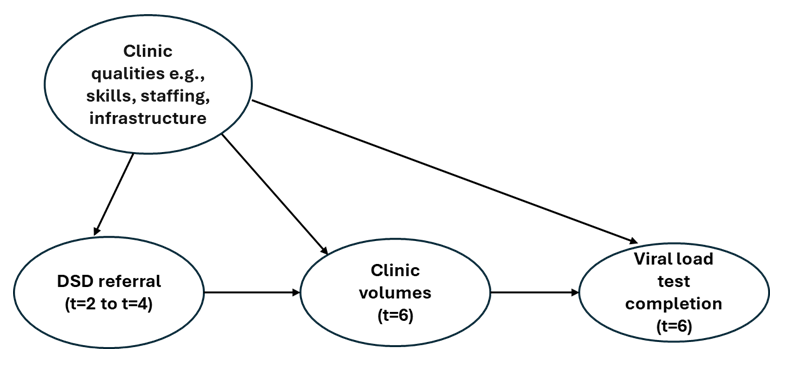

**C**

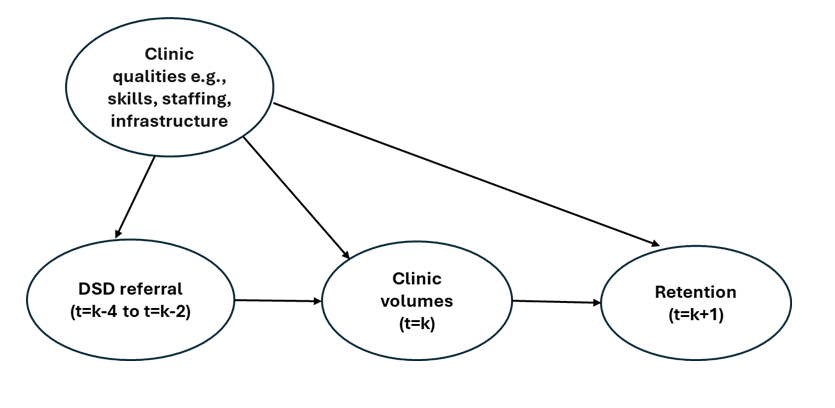
